# Opioid Overdose in Ohio: Comprehensive Analysis of Associated Socioeconomic Factors

**DOI:** 10.1101/19005140

**Authors:** Chihyun Park, Sara Crawford, Rocio Lopez, Anna Seballos, Jean R. Clemenceau, Tyler Coy, Gowtham Atluri, Tae Hyun Hwang

## Abstract

**Objective:** Our study focused on identifying socioeconomic factors associated with death by opioid overdose in Ohio communities at the census tract level.

**Materials and Methods:** A large-scale vital statistic dataset from Ohio Department of Health (ODH) and U.S. Census datasets were used to obtain opioid-related death rate and socioeconomic characteristics for all census tracts in Ohio. Regression analysis was performed to identify the relationships between socioeconomic factors of census tracts and the opioid-related death rate for both urban and rural tracts.

**Results:** In Ohio from 2010-2016, whites, males, and people aged 25-44 had the highest opioid-related death rates. At the census tract level, higher death rates were associated with certain socioeconomic characteristics (e.g. percentage of the census tract population living in urban areas, percentage divorced/separated, percentage of vacant housing units). Predominately rural areas had a different population composition than urban areas, and death rates in rural areas exhibited fewer associations with socioeconomic characteristics.

**Discussion:** Predictive models of opioid-related death rates based on census tract-level characteristics held for urban areas more than rural ones, reflecting the recently observed rural-to-urban geographic shift in opioid-related deaths. Future research is needed to examine the geographic distribution of opioid abuse throughout Ohio and in other states.

**Conclusion:** Regression analysis identified associations between population characteristics and opioid-related death rates of Ohio census tracts. These analyses can help government officials and law official workers prevent, predict and combat opioid abuse at the community level.

## INTRODUCTION

Opioids have been widely used to control and mitigate chronic and postoperative pain. According to data published in the Center for Disease Control and Prevention (CDC) Morbidity and Mortality Weekly Report in 2017, 130 people died each day from opioid-related drug overdoses.[1] In 2017, the U.S. Department of Health and Human Services declared the opioid epidemic a public health emergency.[2] Despite recommendations against the use of opioids, during the past two decades, sharp increases in opioid prescriptions have resulted in opioid abuse and accidental opioid overdoses. The rate of opioid prescriptions written has steadily decreased since 2010, but the rate of drug overdose deaths involving any opioid continues to increase.[3, 4] The increase of such opioid overdose deaths in 2016 was due to the use of heroin and synthetic opioids other than methadone (such as illicitly manufactured fentanyl).[3, 4] While the United States faces the most serious problems with opioid abuse and opioid-related deaths, opioid abuse is an international issue, such as the trafficking of opioid painkillers for non-medical use in Africa.[5]

To address the continuing opioid crisis, it is necessary to identify the characteristics of patients who have used opioids over a long period of time and have died due to overdose. Several studies have aimed to reveal the association between socioeconomic factors and opioid-related mortality. Previous studies have revealed that poverty status and educational attainment could have an effect on drug overdose and subsequent death.[6-10] One recent study investigated the existence of a social gradient in fatal drug overdose cases related to non-prescribed opioids.[11] The researchers found that the mortality rate of opioid and cocaine users from the lowest socioeconomic group was 9.88 times higher than that of their peers of the highest socioeconomic group. One study estimating associations between socioeconomic indicators and opioid-related deaths for Orange County, California found that males, persons aged 45-54, and people identifying as Caucasian had the highest rate of opioid mortality.[12] Additionally, the highest death rates were seen in homeless adults. Another recent study revealed an association between low income neighborhoods and increased risk of receiving an opioid treatment for back pain.[13] Despite the existing research on the associations between socioeconomic factors and opioid-related mortality, all associated socioeconomic factors have not yet been fully elucidated and may vary from region to region, thus calling for more specific studies.

Recently, large-scale data sources related to opioid misuse have become publicly available, although some require Data Use Agreements (DUAs) to access. We used a large-scale vital statistic dataset from the ODH and U.S. Census datasets to perform retrospective and extensive analysis at a statewide and Census tract level. Our study focused on Ohio, as opioid overdose and abuse has become the most urgent public health issue facing Ohio. In 2017, Ohio had the second highest rate of opioid-involved overdose death in the country, at 39.2 deaths per 100,000 persons [14] compared to the national average of 14.9 per 100,000.[1]

The objective of this study is to reveal the socioeconomic factors of communities that are significantly associated with increased opioid overdose-related mortality rates in Ohio at the census tract level. We expect to produce useful analytic results to help government officials and healthcare providers focus their efforts to solve the opioid epidemic.

## METHODS

### Data Description

With the exception of the rural/urban status, which was extracted from the 2010 Census,[15] all data were obtained from the American Community Survey (ACS) for Ohio from 2010-2016.[16] As a result, all variables changed over time using the updated ACS 5-year estimates, with the exception of the percentage rural/urban status which was fixed across all study years using 2010 Census values. For the state of Ohio, we extracted overall population estimates as well as population estimates by gender, age and race. For each census tract, we extracted the percentage of the population with characteristics related to gender, age, race, ethnicity, education, marital status, household occupancy, employment, poverty, income, health insurance, rural/urban status and housing.

All data on opioid-related deaths were extracted from ODH mortality data containing decedent information by opioid poisoning and misuse (2010-2016). We were approved to access limited and identifiable data, completed the DUA, and de-identified decedent data. Access and use of the data was approved by the ODH IRB. From the entire set of vital records containing information on all deaths in Ohio, we selected only opioid-related records. We considered a death opioid-related if the record had a positive value for any of the following nine indicators: Methadone, opiates, prescription opiates, Fentanyl, Fentanyl and Analogues, Carfentanil, designer opioids, commonly prescribed opioids, and/or other opioids. Then, we chose records for analysis which contained positive values for “Opioid Related Death,” “Unintended Death,” “Undetermined death,” and “Drug Poisoning Injury Mechanism.”

### Analysis

We explored trends in the opioid-related death rate, calculated as the number of opioid deaths per 10,000 persons, between 2010 and 2016 by gender, age and race. We then compared the distributions of gender, age and race among the entire Ohio population to the distributions among those opioid fatalities. Chi-square goodness of fit tests were used to determine whether the distribution of characteristics among the opioid fatalities were different than those of the Ohio population.

We then aimed to explore the relationship between the extracted census tract characteristics and the opioid-related death rate among Ohio census tracts using regression analysis. For the purposes of the regression analysis, those census tracts with a population of zero were removed. We first calculated descriptive statistics (mean and standard deviation) for all of the variables considered for the regression analysis. We then explored the unadjusted relationships between each variable and the opioid-related death rate using Poisson regression with the log of the census tract population serving as the offset. We used a deviance adjustment to the scale parameter for the unadjusted analysis due to slight overdispersion. Because the variables selected for the regression analysis displayed a high degree of multicollinearity, we reduced the number of variables for consideration in the multivariable regression model based on variance inflation factors (VIF) < 5. To explore the relative importance of each category, multivariable Poisson regression models with no adjustment to the scale parameter were fit using stepwise model building and an alpha level of 0.05. The final set of variables used in the multivariable model building are listed in Table 3. The regression analysis was performed separately for each year, since we suspected that relationships between the census tract characteristics and the opioid-related death rates may differ over time. The results for each year are presented overall, as well as separately for mostly urban and mostly rural census tracts, because we also suspected that the rural/urban classification would affect the factors associated with the opioid-related death rates.

Results are reported as the rate ratio (RR) and 95% confidence interval (95% CI) and can be understood as the relative risk of death by opioid overdose in the given group, with a confidence interval encompassing the value of 1 signifying nonsignificance.

We used SAS version 9.4 (SAS Institute) to conduct the analyses.

## RESULTS

Opioid-related death rates have increased over time in Ohio during 2010-2016. Rates are highest among males as compared to females (Figure 1A), among persons aged 25-44 years followed by 45-54 years (Figure 1B), and among whites (Figure 1C).

**Figure 1A.**
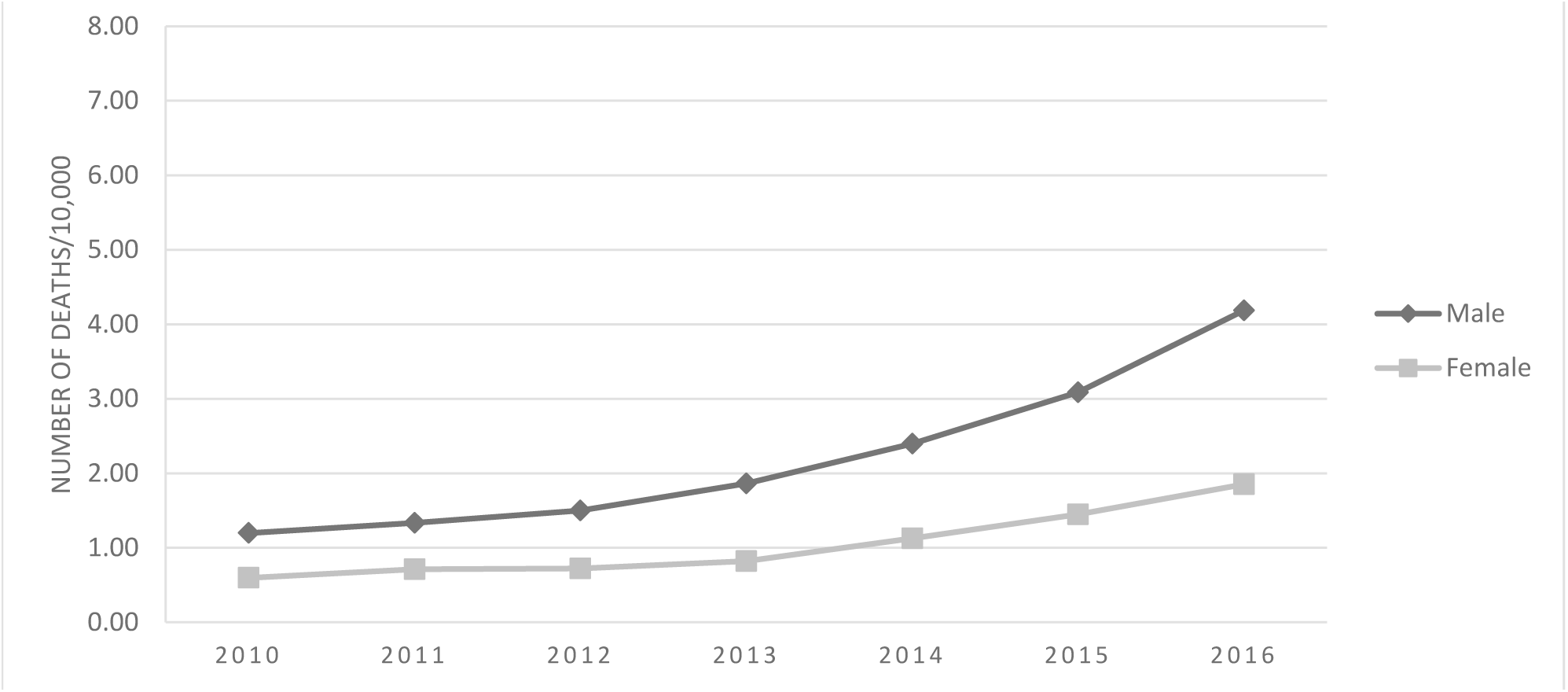
Trends in the opioid-related death rate in Ohio by gender, 2010-2016.

**Figure 1B.**
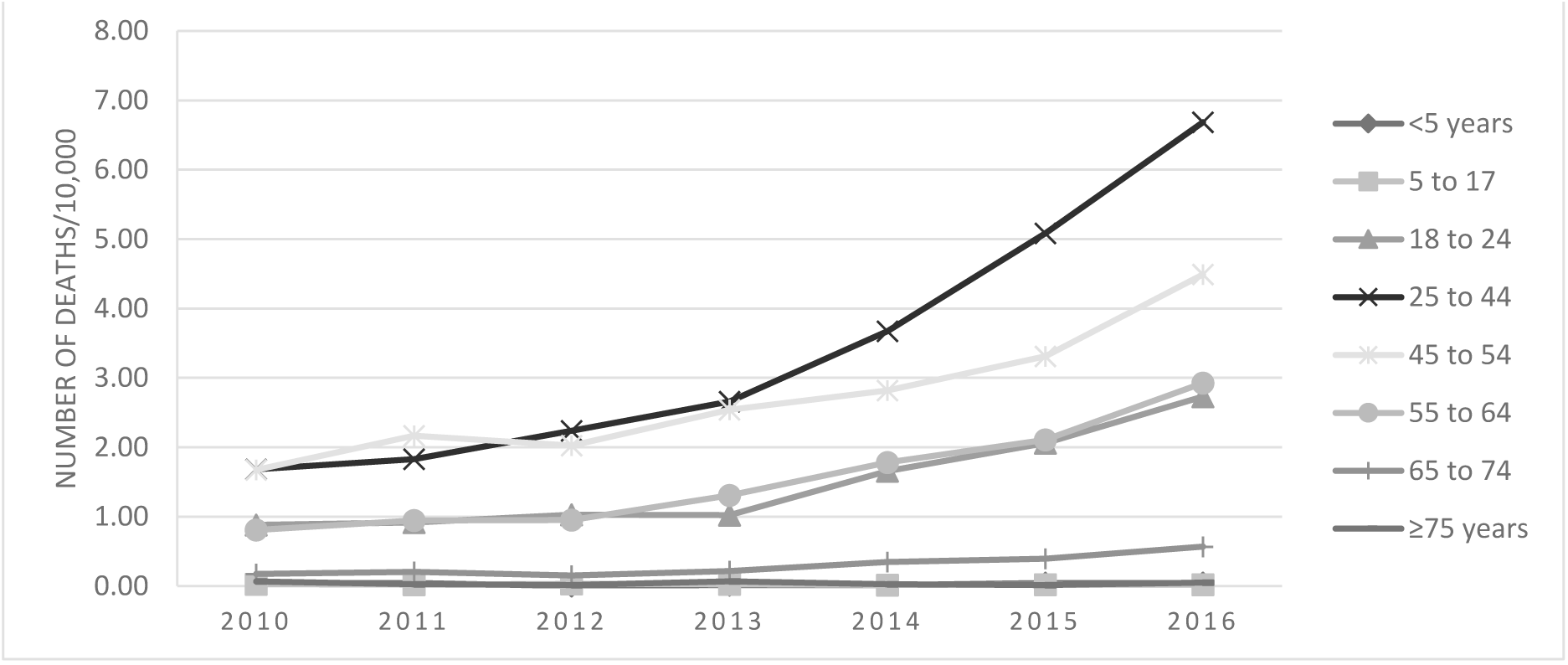
Trends in the opioid-related death rate in Ohio by age group, 2010-2016.

**Figure 1C.**
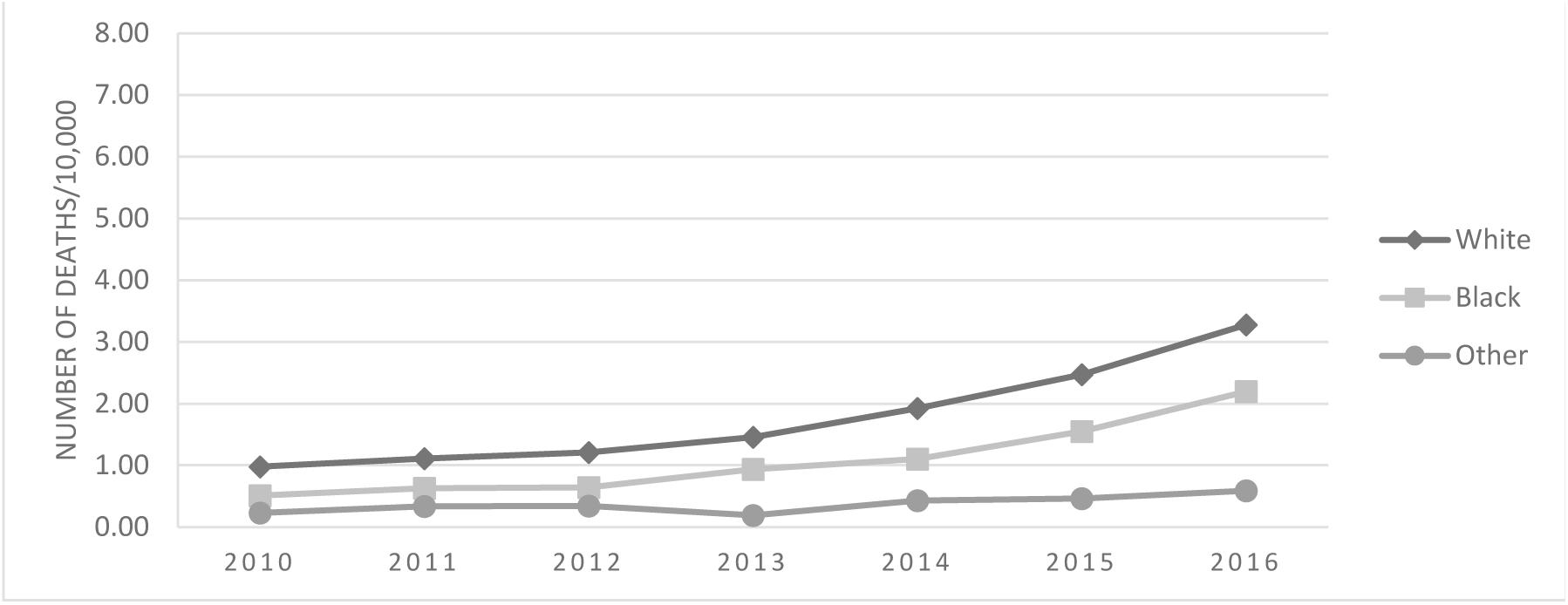
Trends in the opioid-related death rate in Ohio by race, 2010-2016.

These same groups also constitute a higher proportion of opioid deaths compared to that of the general Ohio population, with a higher percentage of males (67.2% vs.48.9%, Figure 2A), individuals 25-44 years and 45-54 years (52.6% vs. 25.1% and 24.3% vs. 14.6%, respectively, Figure 2B), and whites (90.7% vs. 82.8%, Figure 2C).

**Figure 2A.**
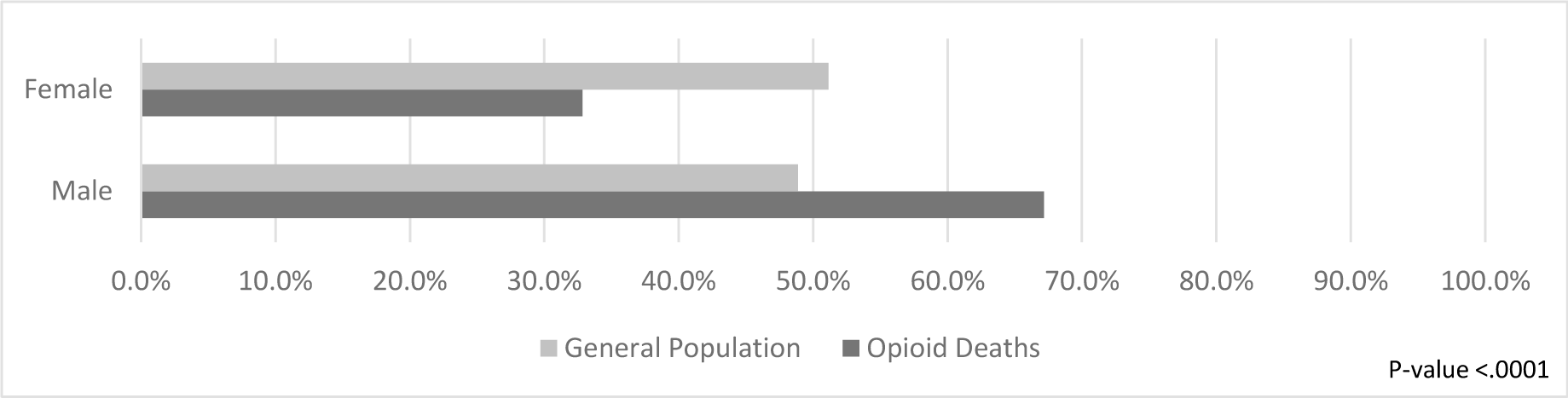
Distribution of gender among the population and among opioid fatalities in Ohio, 2010-2016. (Results combined across years.)

**Figure 2B.**
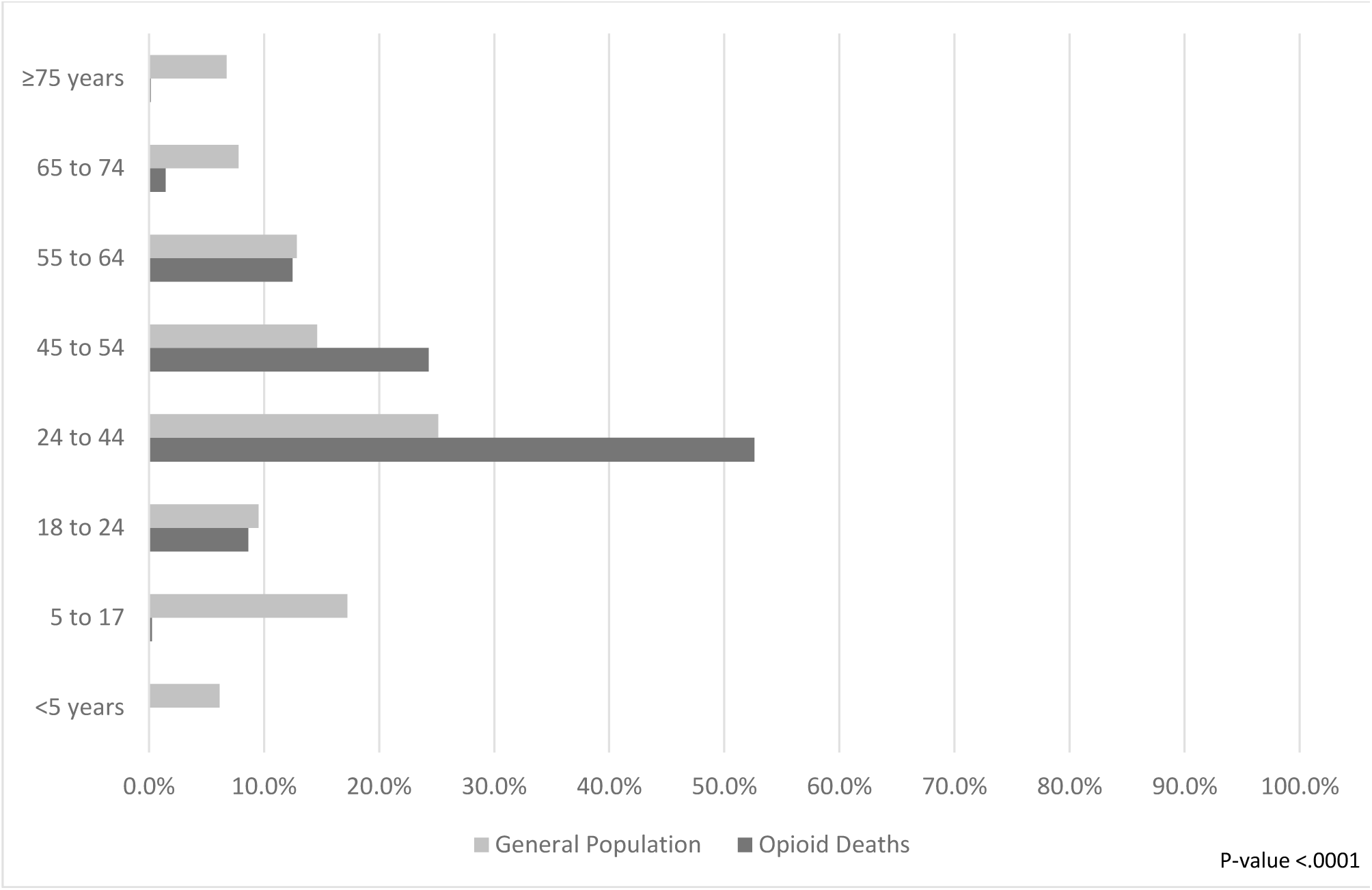
Distribution of age among the population and among opioid fatalities in Ohio, 2010-2016. (Results combined across years.)

**Figure 2C.**
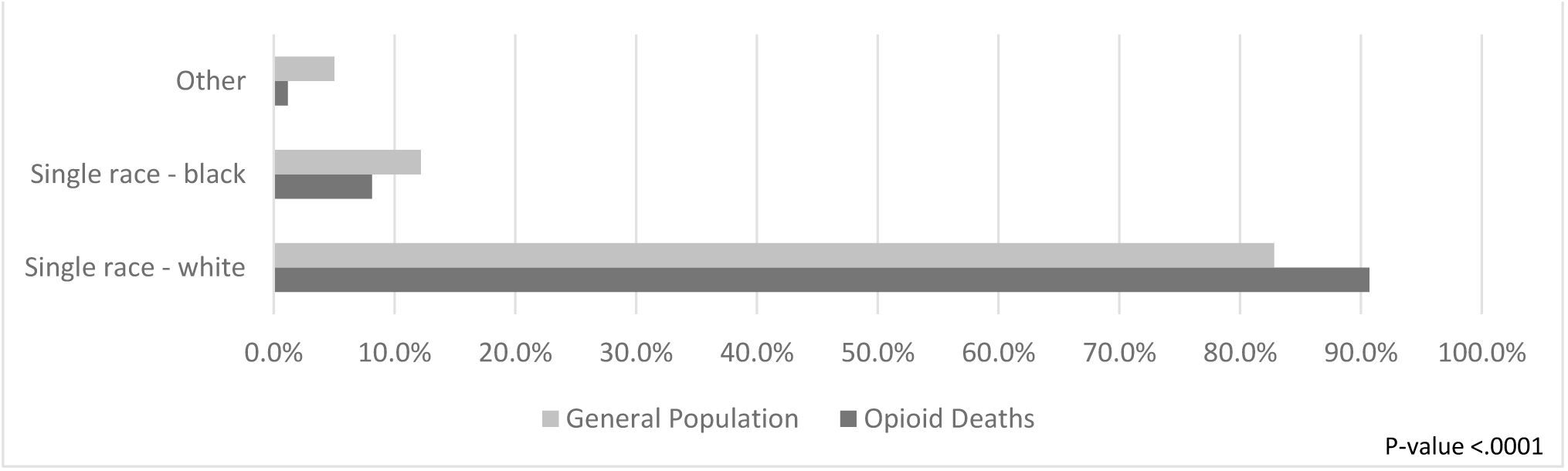
Distribution of race among the population and among opioid fatalities in Ohio, 2010-2016. (Results combined across years.)

In general, the distributions of the census tract characteristics appear similar over time (Supplementary Table 1). On average, the census tract populations have a larger percentage of females, persons aged 25-44 years, whites, persons with at least a high school degree, and persons not married. On average, almost half of households have a single head, about a quarter are living under 150% of the poverty level, and about 80% live in predominately urban areas. The associations between the census tract characteristics and the opioid-related death rate varied across the years, but certain patterns did emerge both prior to adjustment (Supplementary Table 2) and after adjustment for the variables in the multivariable models (Table 1).

**Table 1.**
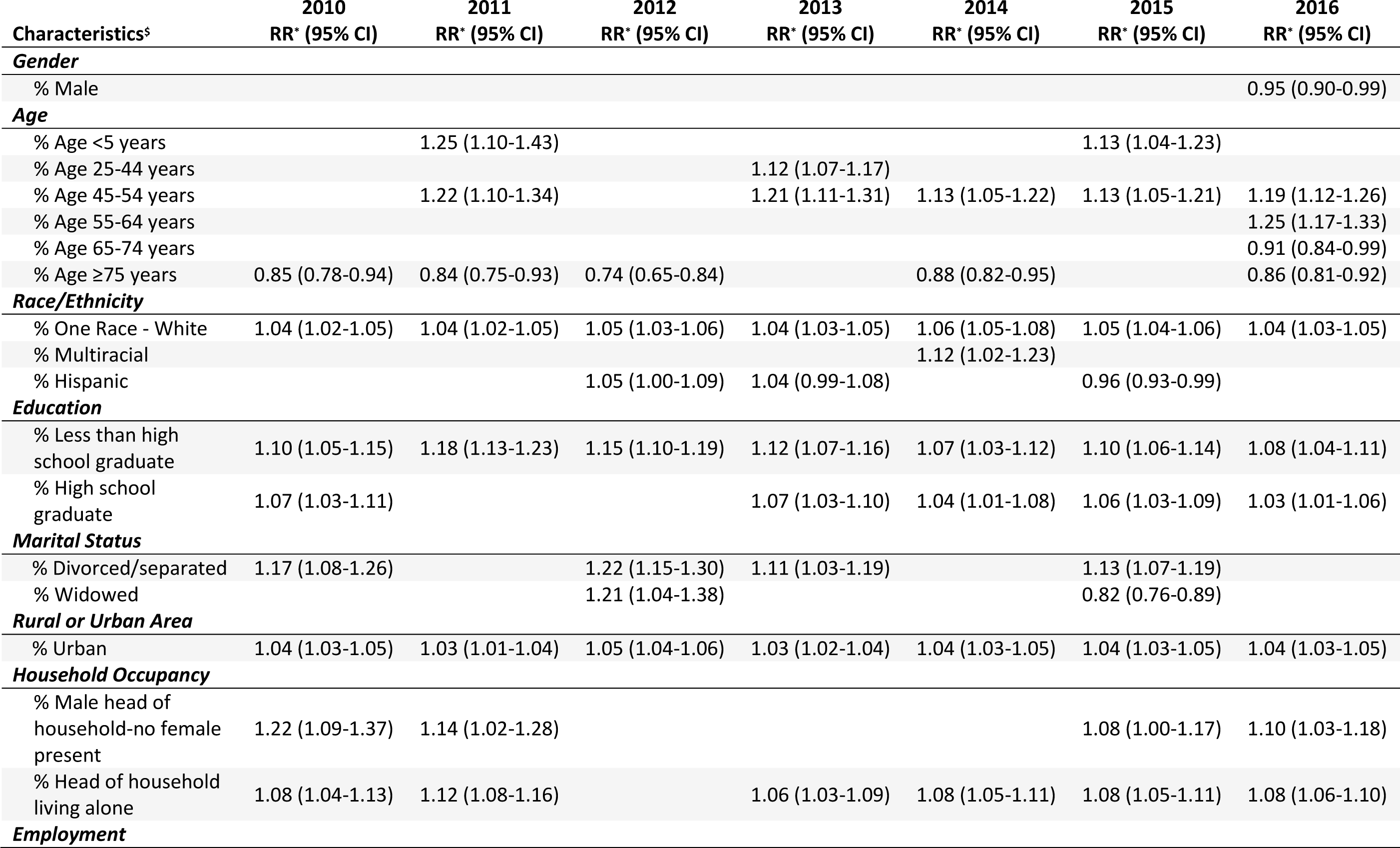
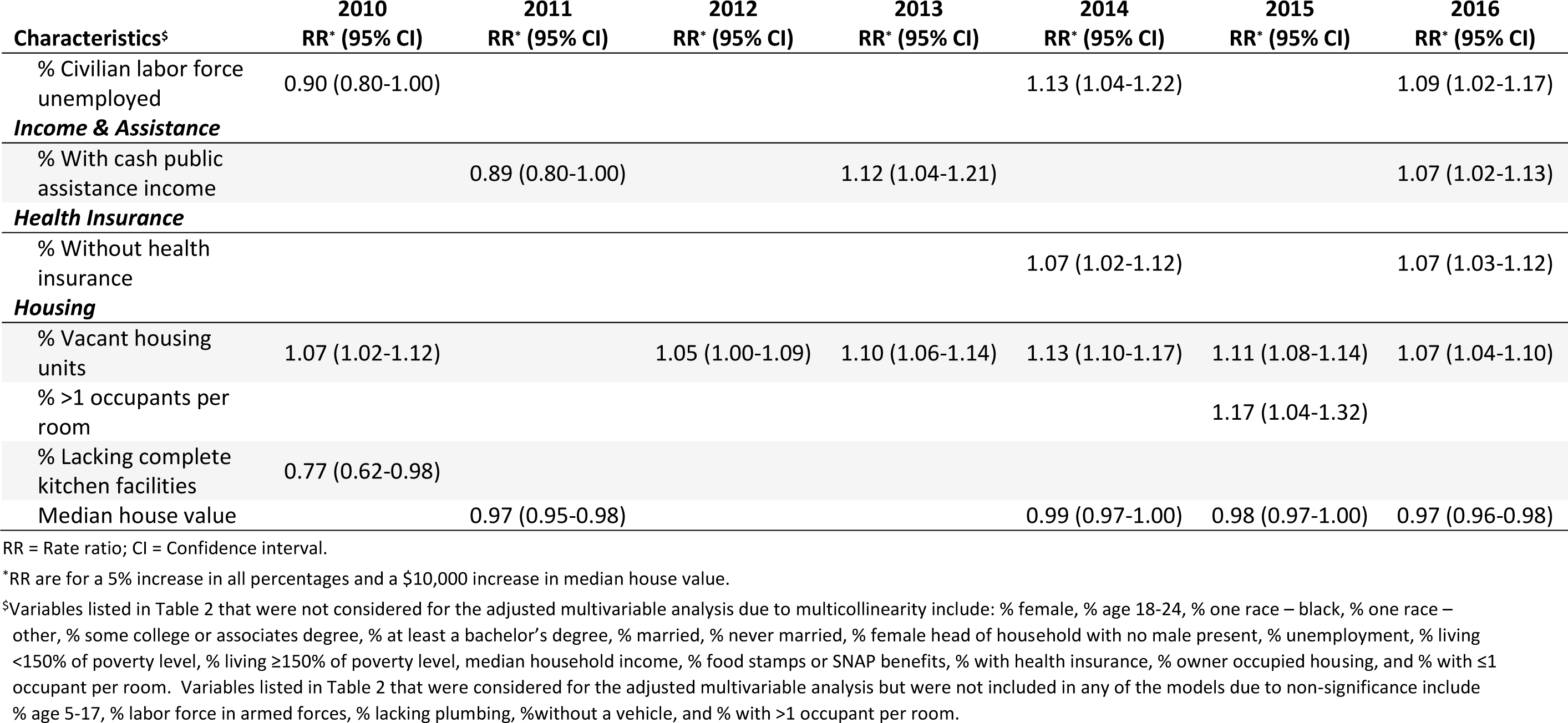
Adjusted associations between census tract characteristics and opioid-related death rates, Ohio, 2010-2016.

After adjustment, an increase in the percentage of the population 54 years or younger was associated with an increase in the death rate. Having an increase in the percentage of the population 65 years and older was associated with a decrease in the death rate. Census tracts with higher opioid-related death rates were also associated with a greater percentage of whites, divorced/separated people, education levels up to high school completion, people living in urban areas, male headed households with no female present, head of households living alone, and vacant housing units. In 2016, for a 5% increase in the percentage of 45-54 year olds, the opioid-related death rate was 1.19 (95% CI=1.12-1.26) times higher. In 2016, for a 5% increase in the percentage of whites, the opioid-related death rate was 1.04 (95% CI=1.03-1.05) times higher; for a 5% increase in the percentage without a high school diploma, 1.08 (95% CI=1.04-1.11); for a 5% increase in the percentage with only a high school diploma, 1.03 (95% CI=1.01-1.06); for a 5% increase in the percentage in urban areas, 1.04 (95% CI=1.03-1.05); for a 5% increase in the percentage of male headed households with no female present, 1.10 (95% CI=1.03-1.18); for a 5% increase in the percentage of head of households living alone, 1.08 (95% CI=1.06-1.10); and for a 5% increase in the percentage of vacant housing units, 1.07 (95% CI=1.04-1.10). A higher median house value was associated with a lower opioid-related death rate; in 2016, the death rate was reduced by a factor of 0.97 (95% CI=0.96-0.97) for every $10,000 increase in the median house value.

Census tracts that were predominantly rural had, on average, a higher percentages of males, whites, high school graduates (versus at least a bachelor’s degree), married persons, and owner-occupied homes, as well as higher median household incomes than compared to predominantly urban census tracts (Supplementary Tables 3A-3B). Predominantly rural census tracts also had, on average, a lower percentage of female headed households with no male present, households headed by persons living alone, unemployed, households below 150% of the poverty level, and households receiving public assistance or using food stamps. We found that the models of the overall population characteristics against the opioid-related death rates showed few significant associations with the predominantly rural areas prior to adjustment, while the results for the predominantly urban areas more closely reflected the overall results (Supplementary Tables 4A-4B). For the rural census tracts after adjustment, few characteristics were significantly associated with changes in the opioid-related death rates. The only factors showing a rather consistent association over time were the percentage of the population aged 25-54 and the percentage divorced/separated, with both resulting in an increase in the death rate for a 5% increase in the characteristic (Table 2A). However, the adjusted results for the urban census tracts were similar to the overall adjusted results (Table 2B).

**Table 2A.**
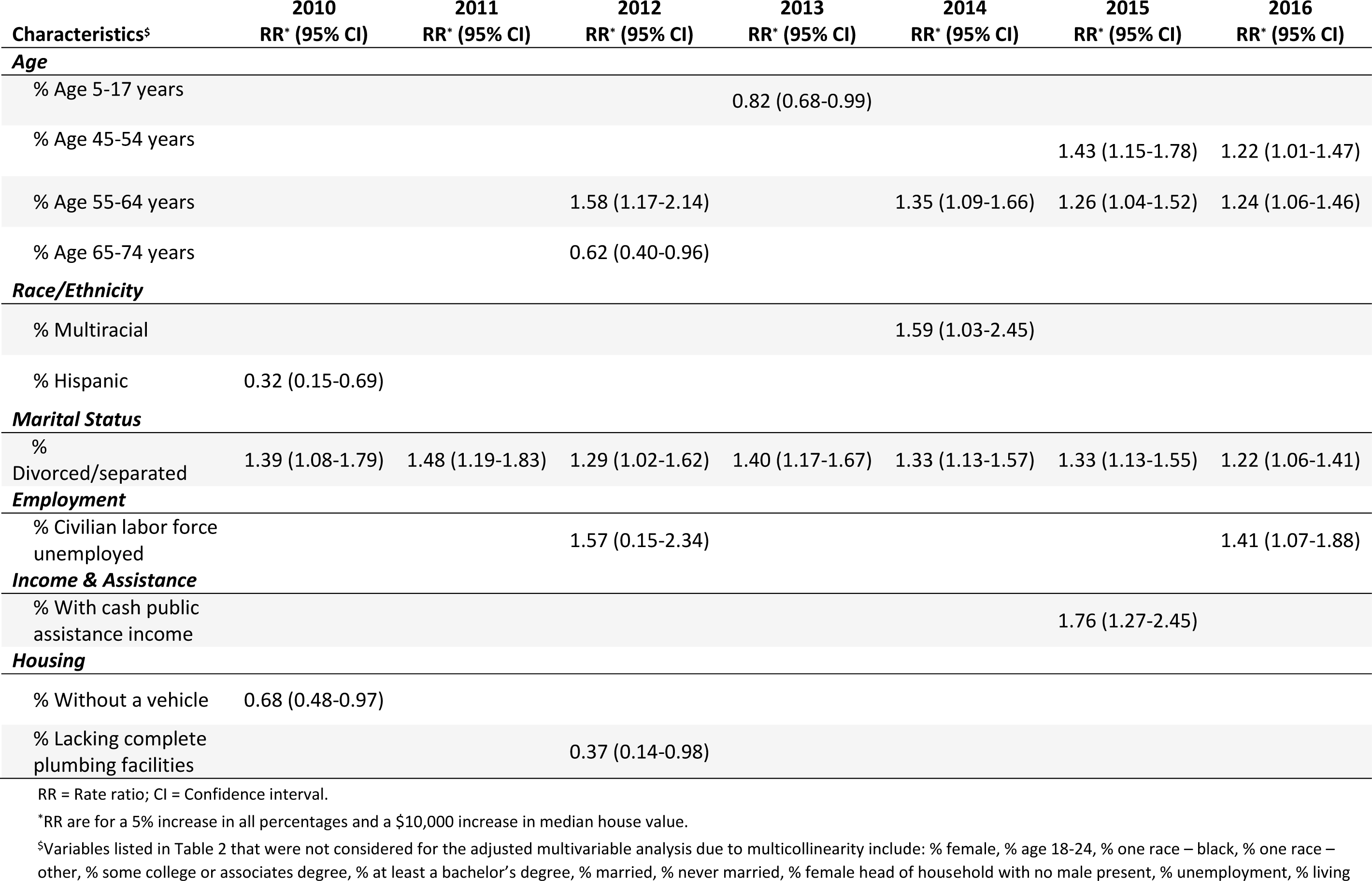
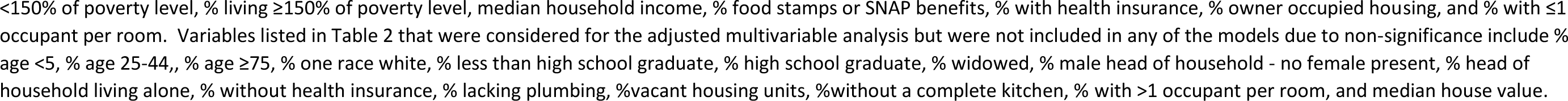
Adjusted associations between census tract characteristics and opioid-related death rates, predominantly rural tracts only, Ohio, 2010-2016.

**Table 2B.**
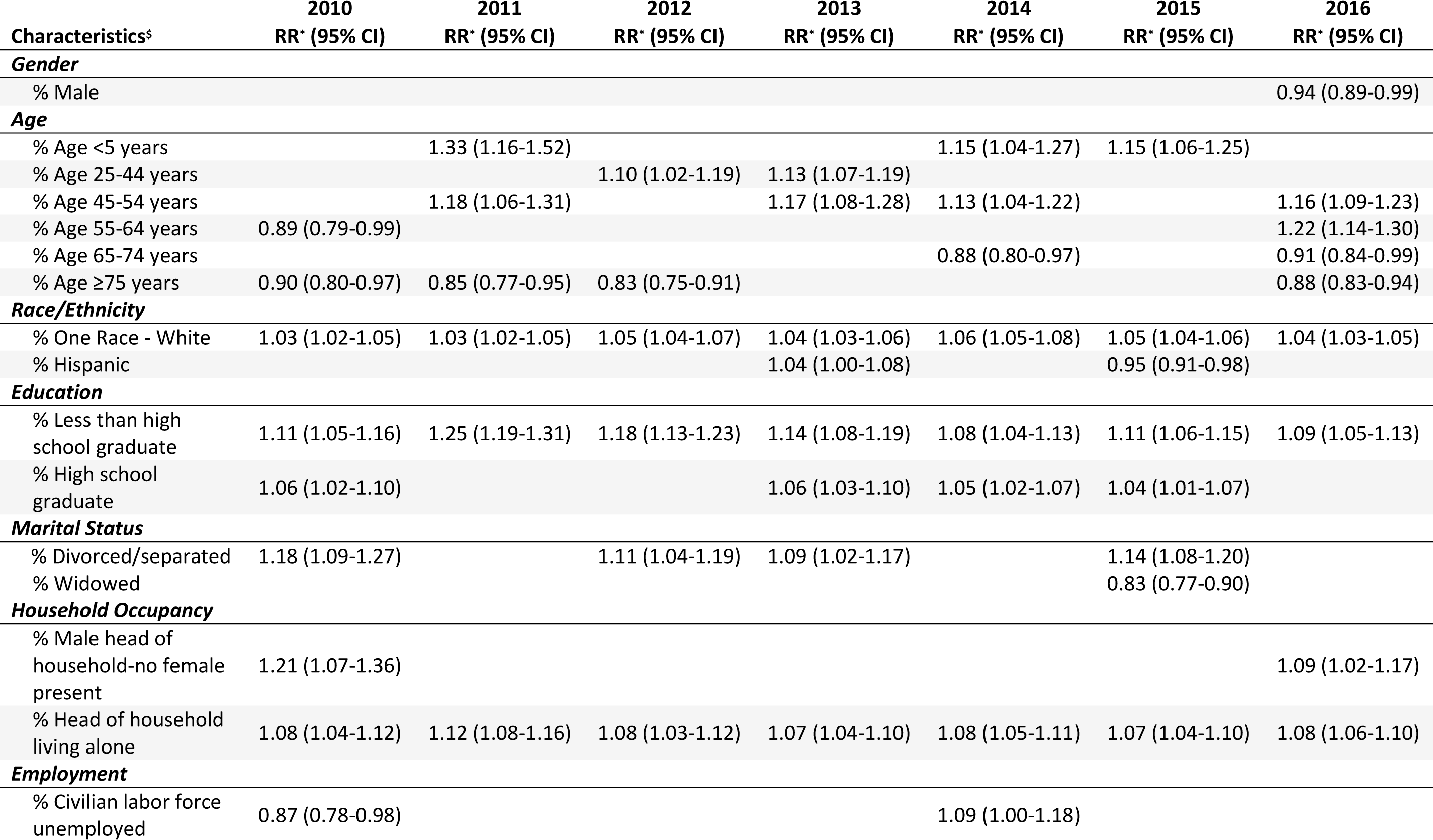
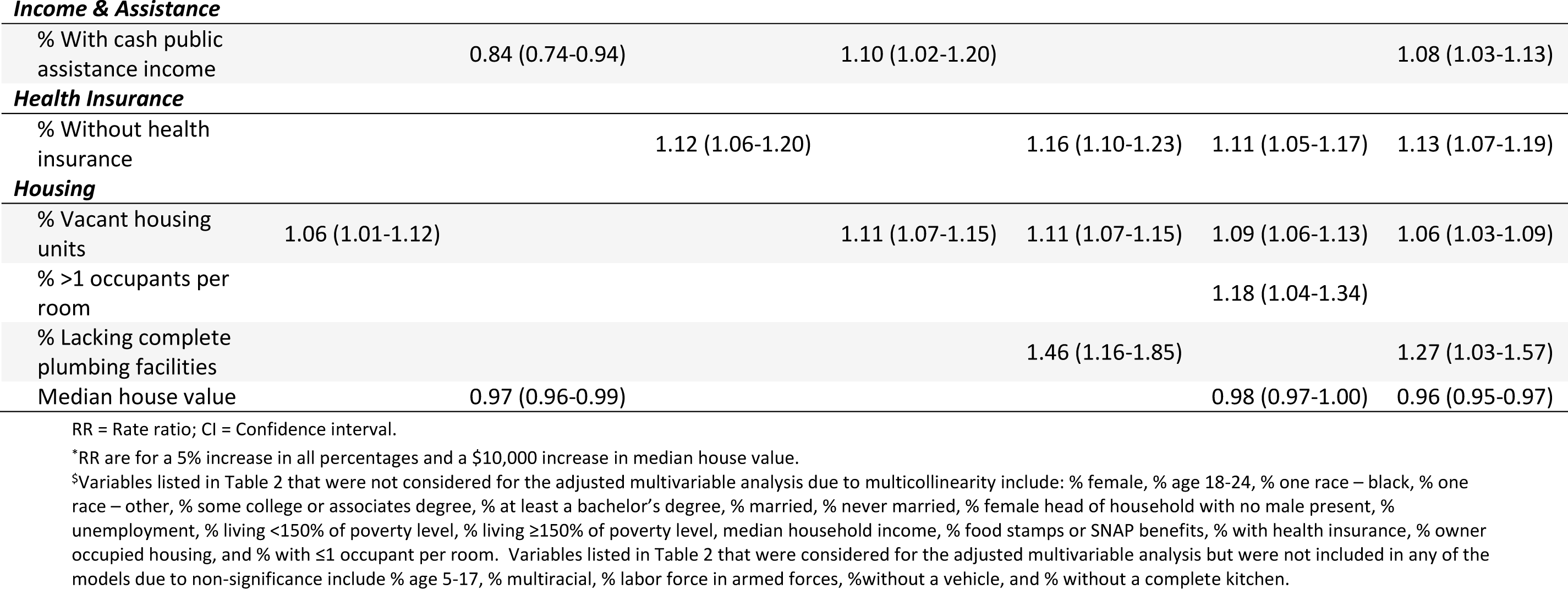
Adjusted associations between census tract characteristics and opioid-related death rates, predominantly urban tracts only, Ohio, 2010-2016.

## DISCUSSION

This study focused on the socioeconomic factors of Ohio census tracts and their association with opioid-overdose related death rates from 2010 to 2016. Regression analysis was performed using mortality data from the ODH and data extracted from the ACS for the state of Ohio.

Analysis of associations between changes in census tract socioeconomic factors and opioid-related death rates in Ohio census tracts found that an increase in the death rate was associated with an increase in the prevalence of the following census tract characteristics: percentage of the population aged 25-55, percentage of whites, percentage of educational attainment less than or equal to high school completion, percentage divorced/separated, percentage living in urban areas, percentage of male-headed households without a female present, percentage of head of households living alone, and the percentage of vacant housing units. A higher median house value was associated with a lower death rate. These findings are interesting as they reflect the increase of drug overdose deaths in urban areas compared to that of rural areas that was observed in 2017.[17] While the increase in narcotic-related death rates for rural areas surpassed that of urban areas between 1999-2004,[18] recent data shows higher death rates in urban counties.[17] Our findings are also consistent with research that found a significant association between higher numbers housing vacancies and higher rates of opioid-related death.[19]

These associations were found to hold for urban census tracts, while rural tracts did not show the same associations, except for the percentage of the population aged 25-54 and the percent that was divorced/separated. Thus, our research shows that the socioeconomic factors that predict opioid-related deaths in urban populations do not hold for rural populations. While this observation may be due to the more homogeneous makeup of rural areas or their smaller sample size, it may also indicate the changing geographical patterns of opioid abuse. Census tract level socioeconomic characteristics other than those that are presented here should be considered in order to predict opioid abuse in rural tracts, in addition to researching which drugs are used more widely in urban versus rural areas.

Limitations of this study include the exclusion of several variables from the multivariable model building, as they exhibited multicollinearity. Additionally, the analysis is based on census-tract level data and shows associations between community characteristics and community death rates and does not explore personal risk factors for opioid-abuse related deaths. Finally, all variables except the information about rural and urban communities came from the ACS and thus were updated on an annual basis, while the rural and urban data came from the 2010 Census data and is fixed for each year analyzed.

We suggest further research to analyze these associations with population data from other states to verify if the model can be expanded beyond Ohio. Additionally, further research on the spatial patterns of opioid-related deaths is needed to explore the incidence of opioid abuse in rural versus urban locations.

## CONCLUSION

The rate of opioid-related deaths has consistently increased in Ohio since 2010. The highest death rates were associated with higher percentages of whites, males, and persons aged 25-54. Our study revealed significant associations using regression analysis between population characteristics of Ohio obtained from the American Community Survey for 2010-2016 and the opioid-related death rate at the census tract level during these years. Census tracts with higher death rates also exhibited higher percentages of the census tract population with educational attainment less than or equal to high school completion, higher percentages of persons divorced/separated, higher percentages in urban areas, higher percentages of male-headed households without a female present, higher percentages with the head of household living alone, and higher percentages of vacant housing units. These patterns found in the overall population did not fit predominately rural populations, but results for predominately urban populations were similar to those of Ohio’s overall population. Future studies should explore these patterns in other states as well as analyze the geographic pattern of opioid misuse to better understand the divergent patterns of opioid use in urban versus rural areas. Our research contributes to greater understanding of predictors and patterns of opioid abuse, and will aid law enforcement agents, emergency health services, and healthcare workers to combat the opioid crisis more effectively.

## Data Availability

Data were acquired from publicly available records from the 2010 U.S. Census and the American Community Survey (2010-2016). Limited-access mortality data were obtained from the Ohio Department of Health via a DUA. Access and use of the data was approved by the ODH IRB.

https://www.census.gov/programs-surveys/acs/data.html

## ACKNOWLEDGMENTS

The authors would like to acknowledge Jesse Schold and Megan Snair of the Center for Populations Health Research of the Lerner Research Institute for their collaboration, Nabhonil Kar for his assistance on the manuscript, and the Ohio Department of Health for access to vital statistics data.

## Contributorship Statement

Conception or design of the work: GA, THH,

Data analysis and interpretation: CP, SC, JRC, RL, TC, AS

Drafting the article: CP, SC, RL, AS, JRC, TC, GA, THH

Critical revision of the article: CP, SC, RL, AS, JRC, TC, GA, THH

Final approval of the version to be published: GA, THH

